# Online cognitive behavioural therapy as an initial treatment for psychological distress in patients waiting for therapy: Study protocol

**DOI:** 10.1101/2023.12.14.23299986

**Authors:** Hayley Guiney, Alison Mahoney, Andie Morgan, Anna Elders, Charlie David, Zahra Howell, David Codyre, Charlene Rapsey, Richie Poulton

## Abstract

**Background:** Wait times for mental health treatment are long. This protocol describes a trial to test whether engaging in a self-directed online cognitive behavioural therapy tool (called ‘Just a Thought’) while waiting for psychological therapy is an acceptable and effective intervention for help-seeking patients. The trial will be conducted within the context of a large psychological service in Auckland, New Zealand.

**Methods:** In this parallel group, pragmatic randomised controlled trial, consenting patients with at least five weeks to wait for their first therapy appointment will be randomly assigned to receive Just a Thought (intervention) or waitlist-as-usual conditions plus generic wellbeing information (control) while they wait. Mental health, general health and wellbeing, and treatment satisfaction will be measured across the first ten weeks of the trial, and at 8 months.

**Outcomes:** The primary outcomes are the acceptability and effectiveness of Just a Thought as an initial treatment. Acceptability will be assessed as participants’ satisfaction, and effectiveness as change in psychological distress over time (relative to control). We hypothesise that participants will be more satisfied with Just a Thought than generic wellbeing information and that those who engage in Just a Thought will show greater reductions in psychological distress than those in the waiting-as-usual group.

**Conclusions:** Online cognitive behavioural therapy is efficacious and has the potential to provide people experiencing psychological distress with fast access to evidence-based psychological treatment. This study will provide insight into its potential as an acceptable and effective pre-treatment for people seeking psychological help.

---

Demand is high for mental health services, and consequently wait times for therapy are long (New Zealand Government Inquiry into Mental Health and Addiction, 2018). Research shows that online cognitive behavioural therapy is an acceptable and efficacious treatment for psychological distress (Andrews et al. 2018), and together with its ready accessibility and scalability, it has the potential to provide faster access to evidence-based psychological treatment. Importantly, online cognitive behavioural therapy does not have to be the sole endeavour of highly motivated patients: it also has the potential to be effectively used as a complement to traditional therapies. Indeed, several regulatory health bodies (e.g., the National Institute of Clinical Excellence (NICE); the Royal Australian and New Zealand College of Psychiatrists) include the provision of complementary online cognitive behavioural therapy as part of their best practice guidelines.

Several randomised controlled trials have tested the effects of providing concurrent in-person healthcare and online cognitive behavioural therapy (for a recent meta-analysis, see Wright, Owen, Richards, et al., 2019), but few have assessed whether providing it as an initial treatment for psychological distress while patients are on the waitlist for therapy has benefits for patients and service providers. One observational study found that patients’ anxiety and depression symptoms improved with use of an online cognitive behavioural therapy programme while they were on the therapy waitlist, and that they continued to improve once their therapy started (Duffy, Enrique, Connell, Connolly, & Richards, 2020). However, without a control group, it remains uncertain whether the observed improvements were due to the intervention itself. Thus, little is known about the effectiveness of providing online cognitive behavioural therapy to patients as soon as they are referred for therapy, and whether such an approach is an acceptable model of care in a primary healthcare setting.

## Objectives

The primary aim of this pragmatic randomised controlled trial is to test the acceptability and effectiveness of an online cognitive behavioural therapy programme (called ‘Just a Thought’) in improving wellbeing and reducing symptoms of psychological distress among patients who are on the waitlist for therapy in a large primary healthcare network, compared with usual waitlist care plus generic wellbeing information. Specifically, we aim to test whether Just a Thought is acceptable to patients as an initial treatment while they wait for therapy, and more effective than waitlist-as-usual conditions plus generic wellbeing information in reducing their psychological distress over the first five to ten weeks since joining the waitlist. We hypothesise that the Just a Thought waitlist intervention will be acceptable to most patients and be more effective in reducing psychological distress than usual waitlist care plus generic wellbeing information. If the intervention is effective, the secondary aim is to explore whether, relative to those in the control group, those in the Just a Thought group demonstrate significantly less demand for therapy services as indexed by lower self-reported need for therapy sessions and fewer therapy sessions provided.

## Ethical review and study registration

This study has been reviewed and approved by the Central Health and Disability Ethics Committee in New Zealand (ref: 2023 FULL 12763) and prospectively registered in the Australia and New Zealand Clinical Trials Registry (ACTRN12623000694617). The trial will be conducted and reported in line with the CONSORT 2010 Statement (Shulz, Altman, Moher, & the CONSORT group, 2010).

## Method

### Design

We will conduct a parallel group superiority randomised controlled trial of Just a Thought versus usual care plus generic wellbeing information for patients on the waitlist for therapy in a primary care network. Participants will be allocated in a 1:1 ratio to the treatment or control groups. The trial was co-designed with practice staff, a local focus group comprising previous patients of the service, and study researchers. We also tailored the design based on findings from an analysis of routinely-collected data from the Just a Thought website to understand common patterns of use and effectiveness for the populations we hope to benefit from this intervention (Guiney, Mahoney, Elders, David, & Poulton, 2023).

### Study setting

The study will be conducted with patients who have been referred by a health practitioner for psychological therapy in a large primary healthcare network that provides psychological services (typically up to six sessions with a therapist) for over forty general practice and urgent care clinics in Auckland, New Zealand (Tāmaki Health). Tāmaki Health uses a patient-centred model of care and serves a culturally and socioeconomically diverse population across Auckland, New Zealand, including areas of relatively high socioeconomic deprivation. The intervention will be administered and study measures collected online. Eligible participants will be recruited via phone invitation.

### Participants and recruitment

Participants will be recruited from the Tāmaki Health waitlist for psychological therapy. Once a patient has their first therapy session scheduled (i.e., they are on the waitlist), appointment co-ordinators will ask them whether they consent to being contacted about possible participation in a research project being run through Tāmaki Health. The study project manager will contact those consenting patients, introduce the study, check eligibility criteria, and invite them to participate.

### Inclusion criteria

Patients on the Tāmaki Health waitlist who are aged 16 years and over, fluent in English, have at least five weeks to wait before their first therapy appointment, have an email address, and have access to the internet can be included in the trial.

### Exclusion criteria

Patients who completed Just a Thought in the past three months or are actively suicidal (assessed via the Columbia Severity Rating Scale, C-SSRS) will be excluded.

### Informed consent procedures

At the first phone contact, the study project manager will provide a brief overview of the study and what will be involved, screen for eligibility, and provide eligible patients with a link to the sign-up webpage. The sign-up webpage will include the full study information sheet and consent form. After reading, participants will be asked to provide their name, year of birth, gender, ethnicity, and contact details, and to tick a box to indicate their informed consent to take part.

### Intervention

Those assigned to the intervention group will be directed to the Just a Thought website, which is a free online tool that offers evidence-based cognitive behavioural therapy courses. Just a Thought is derived from THIS WAY UP, a series of online cognitive behavioural therapy courses designed by a team of clinicians and researchers in Australia, and tested for efficacy in over 30 randomised controlled trials (see https://crufad.org/our-research/). Just a Thought purchased the licence for THIS WAY UP courses but redeveloped the content to ensure equitable accessibility and cultural responsiveness across the New Zealand population, while retaining the evidence-based cognitive behavioural therapy content. The Just a Thought courses that will be available for participants to choose from are the Depression, Generalised Anxiety Disorder, Mixed Depression and Anxiety, and Social Anxiety courses, all of which are broken into six parts, and the Insomnia course, which has four parts. Participants will be instructed to choose the course that is most relevant to them. Each part of the course comprises a narrative story of a fictionalised character who is experiencing a particular type of psychological distress and takes approximately 30 minutes to complete. The character’s story and journey teaches psychoeducational and skill-based cognitive behavioural therapy content to the participant. Each part of the story is supported by a ‘lesson’, which takes the information learned and supports the user to apply it to their own challenges and experiences in a worksheet format. Each ‘lesson’ takes approximately 60 minutes to complete. Each course has extra resources and worksheets that support the user to enhance the skills and techniques outlined. Intervention group participants can go through the each part of the course at their own pace (i.e., no formal time restriction or cessation point), but will be encouraged via regular contact from the study project manager to complete one lesson per week.

### Control group

Participants in the control group will experience waitlist-as-usual conditions plus access to a webpage that provides general wellbeing information developed by the New Zealand Mental Health Foundation called the ‘Five Ways to Wellbeing’. Control group participants can go through the Five Ways to Wellbeing at their own pace (i.e., no formal time restriction or cessation point), but will be encouraged via regular contact from the study project manager to engage with and practice using one ‘Way to Wellbeing’ each week. Waitlist-as-usual conditions at Tāmaki Health comprise contact for the purpose of appointment scheduling, the provision of information about a local peer-led mental health support group, and the provision of emergency mental health service contact information.

## Assignment of interventions

Participants will be randomly allocated in a 1:1 ratio to the treatment or control group by a computer-generated random number sequence (www.random.org). Once participants click to indicate their consent to take part in the study, the sign-up webpage will randomly assign them to a group based on the random number sequence; no person will have any control over the group a particular participant is assigned to. Following randomisation, participants and the researchers will not be blind to group assignment. Participants assigned to the control group will be directed to a webpage with information on the Five Ways to Wellbeing. Participants assigned to the intervention group will be directed to the main Just a Thought webpage, where they will be able to choose from the Depression, Generalised Anxiety Disorder, Mixed Depression and Anxiety, Social Anxiety, and Insomnia courses.

## Strategies to improve adherence to interventions

The study project manager will contact all participants at weeks 2, 3, 4, 5, 6, 8, and 10 to check in with them, and to remind them to complete the regular mental health assessment. Participants can choose whether those regular contacts occur by phone, text, or email.

Participants will be prompted to engage in the online material associated with their condition: those in the control group will be asked if they have been able to use one of the Five Ways to Wellbeing tips in the past week, and those in the intervention group will be asked if they have completed any Just a Thought lessons in the past week. The check-in contacts are designed to act as engagement prompts and not to provide any form of therapy for clients. Practical assistance regarding how to access the mental health assessment or the online content relevant to participants’ group assignment (Just a Thought for the intervention group or Five Ways to Wellbeing for the control group) may be provided. The duration of calls and number of contacts will be recorded to allow post-hoc analyses comparing call/contact time with intervention versus control participants. Participants will also be provided with modest-value vouchers to thank them for their participation in the study: NZD20 upon completing the baseline mental health assessment, and NZD50 for completing the mental health assessment at week 6.

## Outcomes

Figures 1 and 2 illustrate the process and schedule of enrolment, interventions, and assessments. Mental health assessment data will be collected on a regular schedule (see Figure 2) rather than linked to how many Just a Thought lessons the intervention group participants have completed to ensure comparability in time between assessments for the intervention and control groups.

**Figure 1.**
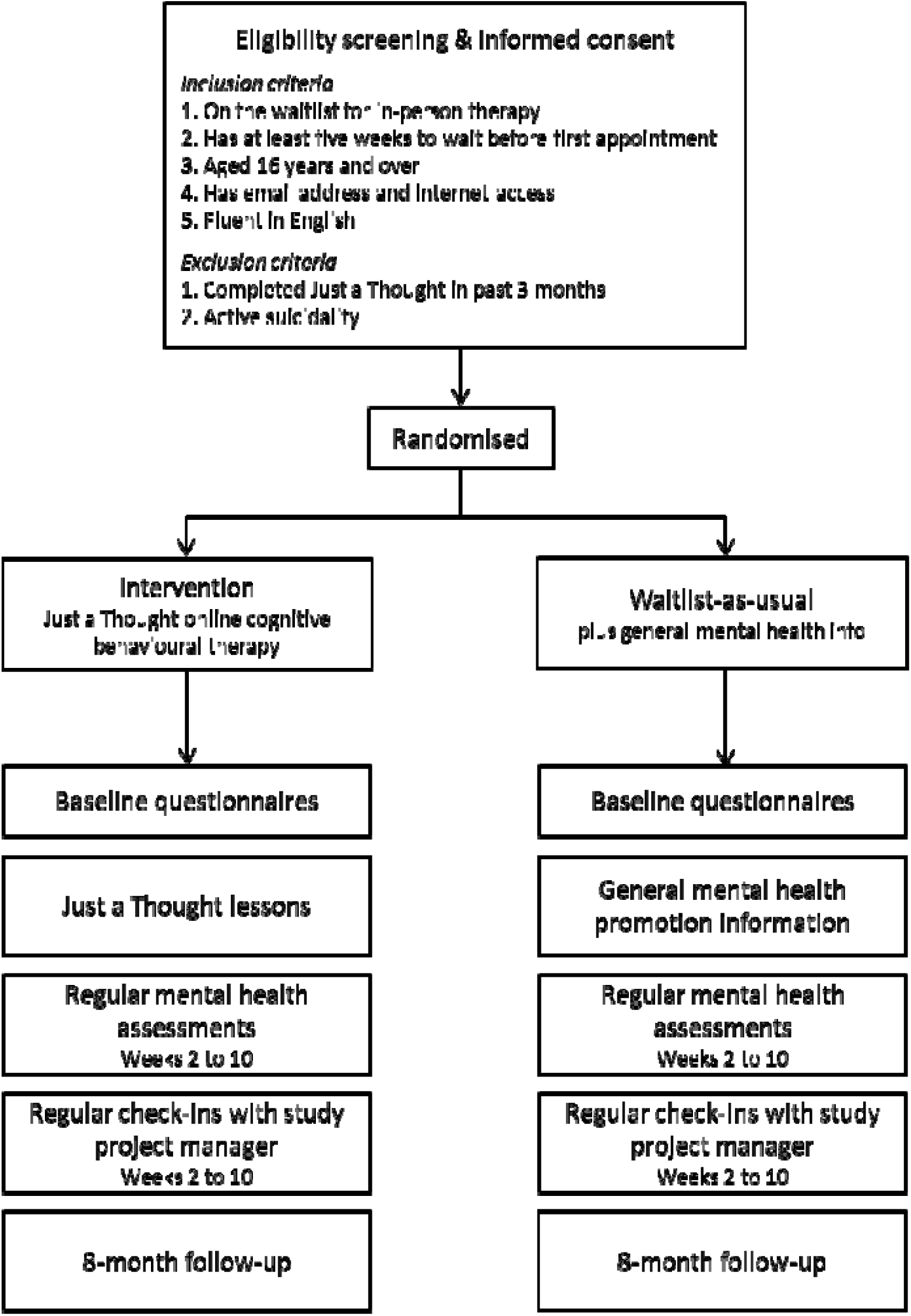
Flowchart showing enrolment and study procedure

**Figure 2.**
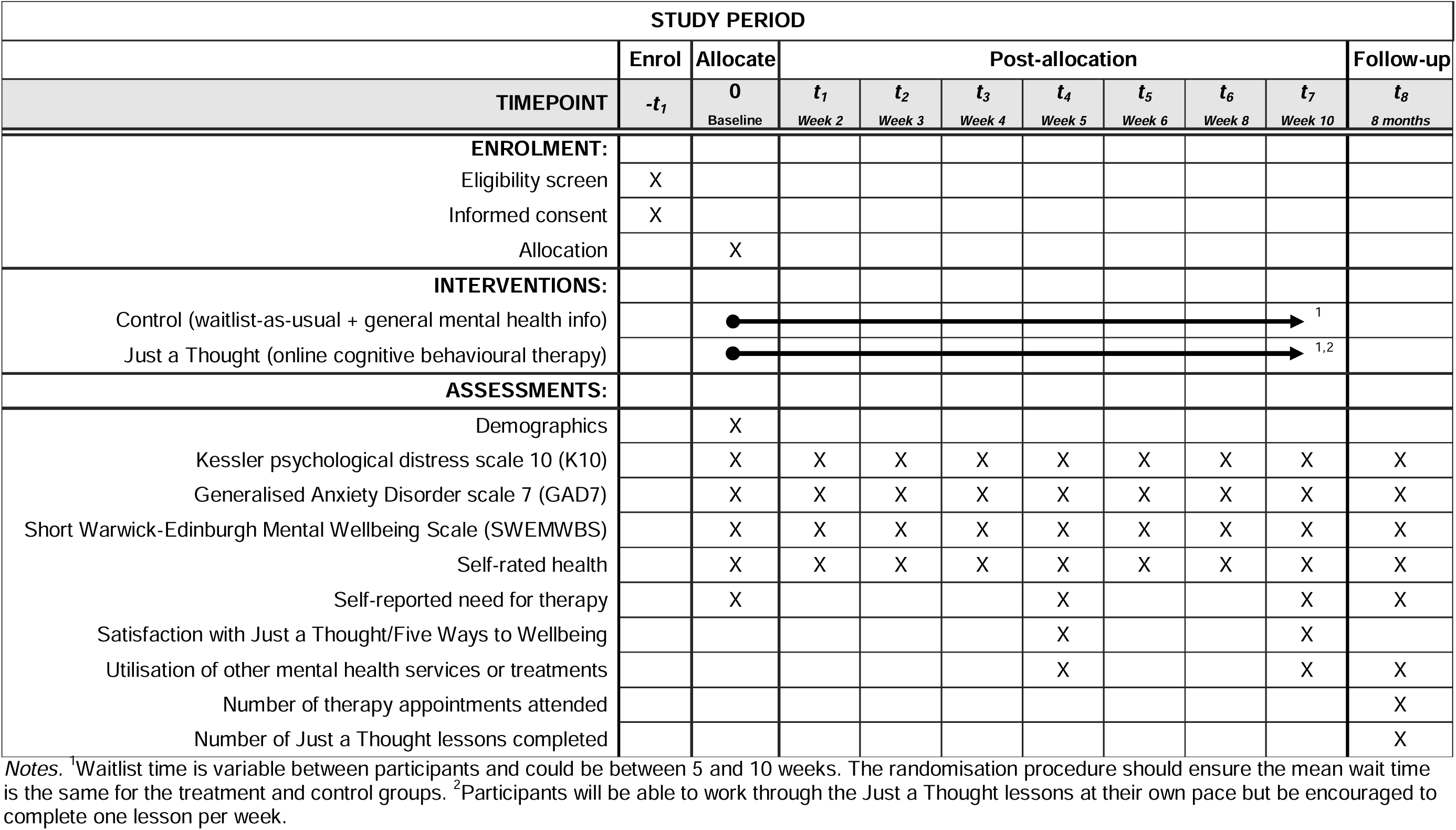
Schedule of enrolment, interventions, and assessments

### Acceptability

Acceptability of Just a Thought as a waitlist intervention will be assessed by asking all participants at week five, ‘On a scale of 1 to 10, how likely would you be to recommend Just a Thought [intervention group]/ the Five Ways to Wellbeing [control group] to a friend or colleague’ (Net Promoter score).

### Effectiveness

All effectiveness measures will be assessed at baseline, at weeks 2, 3, 4, 5, 6, 8, and 10, and at an 8-month follow-up. The primary measure of effectiveness will be participants’ scores on the Kessler-10 psychological distress scale (K10; Kessler et al., 2002). The original form of this 10-item self-report measure assesses non-specific psychological distress over the past 30 days, but for the purposes of the timing in this study, we will modify it to ask about the past two weeks. Possible scores range from 10 to 50, with higher scores indicating greater levels of distress. The K10 has been shown to be an effective method of monitoring changes in psychological distress over the course of other online cognitive behavioural therapy programmes (Sunderland, Wong, Hilvert-Bruce, & Andrews, 2012). The second measure of effectiveness will be scores on the Generalised Anxiety Disorder 7 scale (GAD7; Spitzer, Kroenke, Williams, & Löwe, 2006). This 7-item self-report measure assesses anxiety symptoms over the past two weeks. Possible scores range from 0 to 21, with higher scores indicated greater anxiety. The third measure of effectiveness will be scores on the Short Warwick-Edinburgh Mental Wellbeing Scale (SWEMWBS; Stewart-Brown et al., 2009). This 7-item self-report measure assesses overall mental wellbeing over the past two weeks. Possible scores range from 7 to 35, with higher scores indicating greater mental wellbeing. The fourth measure of effectiveness will be participants’ self-rated health, as assessed by, ‘How would you rate your state of health compared to others your age?’ Participants can respond ‘very good’, ‘good’, ‘fair’, or ‘poor’.

### Secondary outcomes

To assess whether the Just a Thought waitlist intervention is associated with lower demand for therapy services, we will ask participants at baseline, week 5 and 10, and again at the 8-month follow-up to indicate how often in the past 7 days they felt they needed support from a therapist. Participants can respond on a five-point scale ranging from ‘not at all’ to ‘all of the time’. We will also record from the patient management system the number of therapy appointments attended.

### Other measures

At weeks 5 and 10, and at the 8-month follow-up, participants will be asked to report their utilisation of other mental health services or treatments over the past month.

### Safety measures

During the recruitment phone call, the study project manager will assess potential participants’ risk of suicide or serious self-harm via the C-SSRS. The safety response will scale with potential risk severity, ranging from the provision of relevant helpline information to being connected with a senior clinician at Tāmaki Health for in-depth risk assessment and safety plan development. During the study, the regular mental health assessments will include automated risk monitoring. If at any point a participant’s K10 score exceeds 30, indicating very high psychological distress (Andrews & Slade, 2001), they will automatically be sent an email and a text message providing helpline information and encouraging them to seek support. The study website will also include a ‘Get Urgent Help’ button that participants can click to get immediate and relevant helpline information. In addition, any perceived imminent risk identified by the study project manager during interactions with participants will be immediately communicated to a Tāmaki Health clinician via phone call or a secure message to the clinician group for assessment and subsequent action. In the rare situation that a clinician cannot immediately respond, the study project manager will connect the participant to the local crisis team.

### Sample size

Power calculations using observed patterns of change in K10 scores from the New Zealand public’s unguided use of the Just a Thought Depression course indicate that the sample size required to have 80% power to detect a statistically significant time by treatment interaction depends on the number of lessons that those in the intervention group complete. If intervention group participants complete two lessons, a total of between 237 and 345 participants (119 to 173 in each group) is needed. If intervention group participants complete three lessons, a total of between 144 and 188 participants (72 to 94 in each group) will be needed. Given the regular contact by the study manager with participants to check in with them and encourage completion of subsequent Just a Thought lessons, we expect the majority (>50%) of intervention group participants to complete three lessons, and hence have a target total sample size of 200 participants (100 in each group). To allow for a 20% attrition rate, we aim to recruit 240 participants (120 in each group).

### Data collection and management

All data will be collected, encrypted, and stored via the Just a Thought website, which is hosted on Azure Cloud servers in Australia. An audit trail of data accessed and by whom is recorded. Only Just a Thought developers and members of the research team will have access to participant information and identifiable data to monitor participant progress. Only non-identifiable data will be analysed and reported in any outputs, with confidentiality for participants maintained throughout. All non-identifiable data will be stored against a unique identifier.

### Statistical methods

All analyses will be conducted using a standard statistical program, such as Statistical Package for the Social Sciences (SPSS; IBM Corp., USA) or Stata (Stata corp, USA). Descriptive statistics (means and percentages) will be used to assess acceptability of Just a Thought (intervention group) or the Five Ways to Wellbeing (control group). Intention-to-treat mixed models will be used to evaluate the effectiveness of the intervention. First, we will test for a time by treatment interaction in K10 scores, GAD7 scores, SWEMWBS scores, and self-rated health to assess whether change across the first ten weeks of the study is different for those in the treatment versus intervention groups. Second, we will test for a time by treatment interaction in K10 scores, GAD7 scores, SWEMWBS scores, and self-rated health from baseline to post-treatment (8-month follow-up). Mediation and moderation analyses will be used to test how level of engagement in the intervention (i.e., number of lessons completed) and concurrent utilisation of other mental health services during the trial affected the change in scores over time. ANOVAs and/or ordinary least squares regression will be used to test for a between-group difference in participants’ week 4 ratings of how much they feel they need therapy, and the number of therapy sessions participants attended at 8 months follow-up. For all relevant analyses, multiple imputation will be used where appropriate to account for missing data.

## Discussion

Online cognitive behavioural therapy is efficacious (Andrews et al., 2018) and has the potential to be a useful complement to more traditional mental health treatment by providing people experiencing psychological distress with faster access to evidence-based psychological treatment. This novel randomised controlled trial will provide insight into the potential of online cognitive behavioural therapy as an acceptable and effective pre-treatment for patients seeking psychological help from a large primary healthcare network. Using Just a Thought while on the waitlist could lead to greater readiness and motivation for therapy, and act as an orientation to psychological treatment. If the intervention equips patients with new skills that can be further built upon during therapy, improves their wellbeing before they see a therapist, and results in less perceived need (either intensity or duration) for psychological treatment, this work could also be used to help services shorten wait times and meet the huge demand for mental healthcare.

Implementing this trial in the context of a primary healthcare network that serves a culturally diverse population of people from across the socioeconomic spectrum will also provide new information about the acceptability and effectiveness of offering online cognitive behavioural therapy as an initial and immediate-access treatment in a unique clinical, social, and cultural context. Finally, the trial represents an important test of whether Just a Thought, which is based on a Western psychological model and tested mostly with highly educated and socioeconomically advantaged patient groups, can be effectively applied and adapted in a primary mental healthcare setting to meet the needs of a wide group of patients from diverse cultural and socioeconomic backgrounds.

## Author contributions

Guiney and Mahoney led the development of the research design and implementation processes of the trial, and will conduct the analyses. David is the technical lead for the study. Morgan is the study project manager. Rapsey, Elders, Codyre, and Howell are the primary clinical staff advising and assisting with the trial. Poulton provided oversight and advice on the trial until July 2023, when Rapsey took over that role.

## Data Availability

This is a study protocol and hence no data are associated with this manuscript

